# The shared and disorder-specific resting state functional connectivity associated with emotion regulation between ADHD and bipolar disorder

**DOI:** 10.1101/2024.03.25.24304833

**Authors:** Qian Zhuang, Chuxian Xu, Nuo Xu, Tongtong Wang, Matthew Lock, Shuaiyu Chen

## Abstract

**Background:** Emotion regulation deficits have been frequently observed between ADHD (Attention-deficit/hyperactivity disorder, ADHD) and BD (bipolar disorder, BD) adults, however, common and distinguishable alterations in functional connectivity during rest remain poorly understood.

**Objectives:** The current study was performed to determine the shared and disorder-specific resting state functional connectivity (rsFC) features within the proposed hierarchical emotion regulation model.

**Method:** The whole-brain seed-to-voxel functional connectivity analyses were performed on the neuroimaging data from the OpenFMRI project included 40 ADHD adults, 49 BD adults, as well as 49 age and gender matched healthy controls.

**Results:** Our findings showed significantly shared and disorder-specific rsFC circuits mainly linking to the processes of emotion perception and interception during emotion regulation but not response inhibition or executive control. Such as, the results found significantly enhanced functional connectivity strength between inferior occipital gyrus (IOG)-lingual/fusiform gyrus as well as superior parietal lobule (SPL)-insula circuits in ADHD group compared to healthy group (HG) and BD group, which suggests impaired emotional facial perception and interoceptive processing in ADHD than BD adults. In addition, the connectivity between precuneus and fusiform was significantly lower in both of ADHD and BD group compared to HG and no significant difference was found between patient groups. This indicates similar neural patterns underlying the impairment in emotion classification and emotional experience for ADHD and BD adults.

**Conclusions:** Together these findings can promote the understanding of common and disorder-specific neural mechanism underlying emotion dysregulations and facilitate neuroimaging-based clinical evaluation for ADHD and BD adults.

## 1. Introduction

ADHD (Attention-deficit/hyperactivity disorder, ADHD) and BD (bipolar disorder,BD) are common mental disorders with a higher degree of comorbidity, that nearly one in thirteen ADHD adults was diagnosed with BD, and one in six BD adults with ADHD (Schiweck et al., 2021). While ADHD is defined as persisting symptoms of age-inappropriate inattention and/or hyperactivity/impulsivity in DSM-5 (American Psychiatric Association, 2013), the cardinal feature of BD is a lifetime occurrence of depressive or manic episodes. There are many overlapping features and diagnostic criteria between ADHD and BD, including hyperactivity, distractibility, lack of inhibition and irritability, etc (Brus et al., 2014; Comparelli et al., 2022), although some distinctions in symptoms could help to distinguish ADHD from BD. In addition, consistent evidence reveals common risk genetic loci (O’Connell et al., 2021; van Hulzen et al., 2017) and strong familiar association between ADHD and BD (Khoury et al., 2023; Skirrow et al., 2012), which makes the clinical delineation and disorder diagnostic more challenging. Therefore, a better understanding of the common and distinct neural features between ADHD and BD may have great clinical implications.

A growing number of studies have been conducted to identify the common and distinguishable cognitive indices or neural biomarker between ADHD and BD group (Passarotti and Pavuluri, 2011; Pavuluri et al., 2009; Xie et al., 2022; Yep et al., 2018), especially in the dimension of emotion regulation (ER). Emotion regulation, refers to goal-directed processes to modulate the duration, intensity and frequency of experienced positive or negative emotions with strategies (Boemo et al., 2022). While previous findings holding the dual opposing system suggest that ER is mainly linked to the prefrontal-amygdala circuits (Banks et al., 2007; Johnstone et al., 2007) and the regions included dorsolateral and ventrolateral prefrontal cortices (DLPFC and VLPFC), orbitofrontal cortex (OFC) and amygdala showed significant alterations in ADHD and BD (Bigot et al., 2020; Furlong et al., 2021; Kebets et al., 2021; Simonetti et al., 2022; Yep et al., 2018), emerging evidence from recent models and meta-analyses have proposed that ER is relying on multiple neural systems which are implicated in the interaction between emotion perception, generation and regulation (Messina et al., 2015; Morawetz et al., 2017; Morawetz et al., 2020). Particularly, one meta-analytic study based on convergent brain activation patterns involved in ER have identified four large-scale brain networks related to different psychological processes of emotion regulation, wherein the first two are mainly associated with executive control and response inhibition during ER, and the second two are primarily associated with emotion generation, perception, and physiological processes (Morawetz et al., 2020).

In contrast, deficits in ER can be defined as emotion dysregulation (ED), which reflects maladaptive ways during emotion experiences and responses, such as difficulties in emotion understanding or limited access to context-appropriate emotional regulatory strategies (Gratz and Roemer, 2004). Growing evidence from both human behavioral and neuroimaging studies suggest that ED is a trans-diagnostic risk factor as well as the treatment and intervention foci across many mental disorders including ADHD and BD, although it is not included as a formal diagnostic criterion in DSM-5 (American Psychiatric Association, 2013; Carmassi et al., 2022; Kebets et al., 2021; Lenzi et al., 2018). Such as, consistent findings demonstrated that both ADHD and BD patients reported higher ED level and preferred to adopt more maladaptive ER strategies compared to healthy controls (Beheshti et al., 2020; De Prisco et al., 2022; Minò, 2022; Miola et al., 2022; Oliva et al., 2023; Shaw et al., 2014; Soler-Gutiérrez et al., 2023). On the other hand, some other studies show that while BD group exhibited higher scores on affective instability than ADHD group (Richard-Lepouriel et al., 2016; Sesso et al., 2021), ADHD group scored higher on emotional intensity (Richard-Lepouriel et al., 2016; Torrente et al., 2017). More specifically, the studies on the neural level also reveal some common and distinct brain areas involved in emotion processing and regulation between ADHD and BD group (Guan et al., 2023; Passarotti et al., 2010a, b; Phillips and Swartz, 2014). Such as, ADHD group showed decreased VLPFC activation relative to healthy group (HG) and BD group during emotion processing, whereas both patient groups exhibited greater activation in DLPFC and parietal cortex relative to HG (Passarotti et al., 2010a).

Notably, numerous studies have been conducted to detect the resting state functional connectivity profiles between ADHD and BD (Barttfeld et al., 2014; Lopez-Larson et al., 2009; Son et al., 2017). For example, while one study shows enhanced functional connectivity in frontotemporal and fronto-occipital networks for both ADHD and BD groups (Barttfeld et al., 2014), another one finds increased connectivity between the affective network node (i.e. amygdala) and ventral attention network node (i.e. temporoparietal junction) in BD group compared to ADHD group (Son et al., 2017). However, while recent meta-analysis has identified four interacting large-scale brain networks associated with different psychological processes during emotion regulation which has been proved to be with high reliability (Berboth et al., 2021; Morawetz et al., 2020), the common and distinguishable resting state functional connectivity (rsFC) patterns between ADHD and BD within this hierarchical framework of emotion regulation model still remains to be unclear.

To address this issue, the present study was conducted to determine the shared and disorder-specific neural features in terms of resting state functional connectivity associated with emotion regulation between ADHD and BD group using the shared open dataset from the OpenFMRI project (http://openfmri.org; Gorgolewski et al., 2017). In addition, to investigate whether the rsFC strength between ADHD and BD group were associated with the individual differences on executive functions, we additionally examined the correlation between functional connectivity strength and cognitive behavioral performance assessed by stop signal task (SST), Balloon analog risk task (BART) and task-switching in each group. Based on previous findings (Lopez-Larson et al., 2009; Passarotti et al., 2010a; Son et al., 2017; Xie et al., 2022), we expected to see common altered rsFC patterns related to the domain of executive control and response inhibition between BD and ADHD group as well as significant differences in circuits associated with emotion generation, perception and memory. Especially, we hypothesized that there was significant correlation between the cognitive behavioral performance and rsFC circuits linking to executive control or response inhibition.

## 2. Material and Methods Participants

The neuroimaging data used in the present study was from the Consortium for Neuropsychiatric Phenomics dataset which was shared via the OpenFMRI project (http://openfmri.org; Gorgolewski et al., 2017). In detail, the participants (age from 21 to 50) were comprised of n=40 ADHD, n=49 bipolar disorder, and n=50 schizophrenia as well as n=130 healthy controls and were recruited by the community advertisements and outreach to the local clinics and online portals respectively. All of them received at least 8 years formal education and wrote the informed consent before experiments. The exclusion criteria for MRI scanning are as following: left-handedness, insufficient vision, mood-altering medication on test day, history of significant medical illness, and contraindications for MRI (including pregnancy).

### Resting state fMRI data acquisition

Neuroimaging data were collected on 3T Siemens Trio scanners. The resting fMRI data scanning last 304 seconds, during which participants were asked to keep eyes open and remain to be relax. Finally, for each participant, 152 brain volumes were obtained using a T2*-weighted echoplanar imaging (EPI) sequence with acquisition parameters as following: slice thickness=4 mm, 34 slices, TR=2 s, TE=30 ms, flip angle=90°, matrix 64 × 64, FOV=192 mm, oblique slice orientation. In addition, high-resolution whole brain T1-weighted images were collected with following parameters: slice thickness=1 mm, 176 slices, TR=1.9 s, TE=2.26 ms, matrix =256 × 256, FOV=250 mm, sagittal plane.

### MRI data analysis

#### MRI data preprocessing

Given the main aim of the present study was to explore the common and distinguishable rsFC circuits between ADHD and BD groups associated with emotion regulation in a hierarchical organization, only participants from ADHD, BD and healthy groups were included in the data analyses. In addition, given the number of subjects in healthy group largely differed from the other two groups, n=49 participants were randomly selected from the healthy group for whom age and gender were matched between three groups (age: HG(28 male): Mean _age_±SEM = 32.57±1.36, ADHD (21 male): Mean _age_±SEM = 32.05± 1.65, BD (28 male): Mean _age_±SEM = 35.29±1.29). MRI data were preprocessed by SPM12 (Wellcome Trust Center of Neuroimaging, University College London, London, United Kingdom) and CONN (version: 21.a; Whitfield-Gabrieli and Nieto-Castanon, 2012) with standard pipelines. The first 4 volumes were discarded to gain magnet-steady data. Preprocessing for the remained volumes included: (1) spatial realignment using realignment and unwarp; (2) temporal realignment using slice timing; (3) outlier detection using ART-based identification of outlier scans; (4) coregistration; (5) spatial normalization; (6) smoothing using a Gaussian kernel with full-width at half-maximum (FWHM) of 6mm. For spatial normalization, T1-weighted structural images were segmented into gray matter, white matter, and cerebrospinal fluid and the transformation information were used to normalize the functional images to Montreal Neurological Institute (MNI) standard space.

### Motion artifacts control

For head-motion control, 10 subjects (3 subjects from HG, 5 subjects from BD group and 2 subjects from ADHD group) were excluded from further analyzing due to excessive head-motion (> 2.5mm and 2.5 degree during whole scanning). In addition, mean-frame-wise displacement (FD, Power et al., 2012) was calculated for the remaining subjects and another 15 subjects (4 subjects from healthy group, 8 subjects from BD group and 3 subjects from ADHD group) with mean FD value > 0.3mm were excluded. There are no significant differences in the mean FD value among the three groups (*F*_(2,110)_ = 0.622, *p* = 0.539). Finally, 42 (24 male) subjects in the healthy group, 36 subjects (20 male) in the BD group and 35 subjects (18 male) in the ADHD group were included in the final statistical data analyzing.

### Functional connectivity analysis

To detect the shared and disorder-specific rsFC circuits between ADHD and BD individuals during the emotion perception, generation and regulation, the whole-brain seed-to-voxel functional connectivity analysis was performed with CONN (Whitfield-Gabrieli and Nieto-Castanon, 2012). In accordance with recent meta-analytic study (Morawetz et al., 2020), 36 ROIs (regions of interest) from 4 emotional regulation networks (ERNs) were determined as seeds (see **Figure 1**), which were defined as 4mm spheres centered on the peak coordinates of each region from the meta-analyze (Morawetz et al., 2020). Thereinto, the ERN1 and ERN2, included 10 and 9 ROIs respectively, are mainly located at fronto-parietal regions and linked to response inhibition, and executive control. While the ERN3 included 8 ROIs are mainly located at subcortical regions and related to memory, emotion perception and generation, the ERN4 included 9 ROIs are mainly related to interoception, body action and awareness and might serve as a hub between generating emotional responses (ERN3) and regulating these responses (ERN1 and ERN2). Furthermore, given previous studies have showed consistent gender differences on emotion regulation and associated brain functional and structural abnormalities in both ADHD and BD group (Jiang et al., 2021; Menculini et al., 2022; Mowlem et al., 2019; Seymour et al., 2017), exploratory analyses examining the gender effect on the hierarchical functional network between ADHD and BD group were conducted. Then, the group difference as well as the group X gender interaction effects on emotional regulation were examined by one-way ANOVAs (Group: HG vs. BD vs. ADHD) and 3*2 full factor ANOVAS (Group: HG vs. BD vs. ADHD; Gender: female vs. male) respectively.

**Fig. 1.**
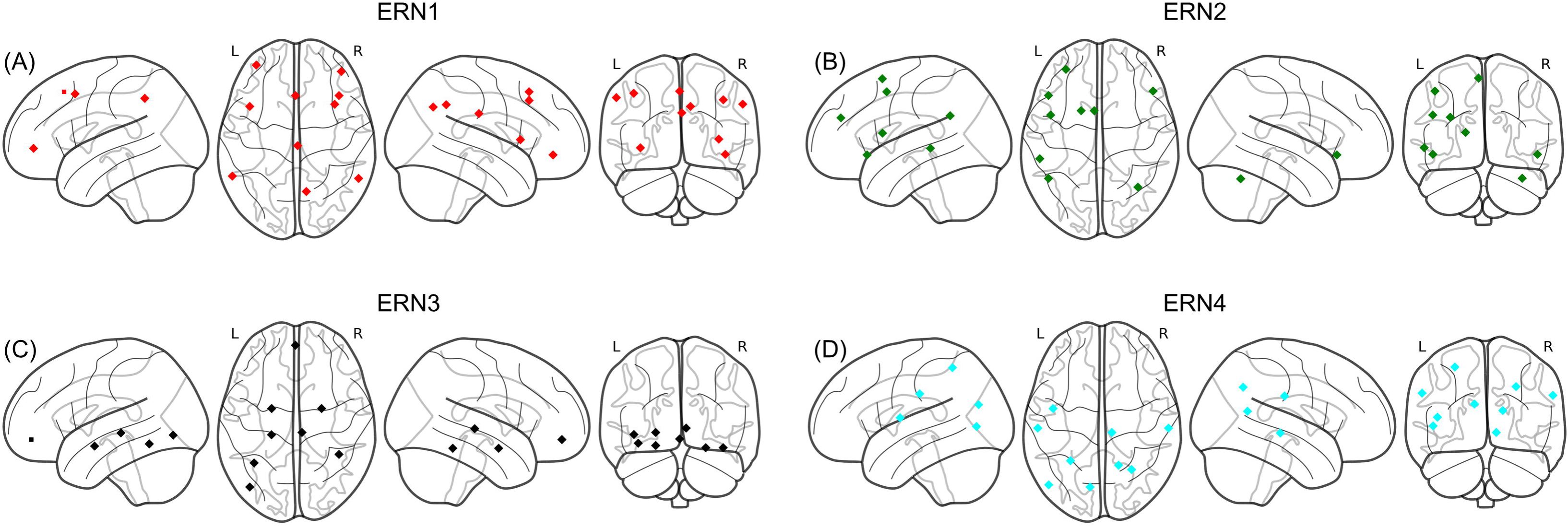
N=36 ROIs (regions of interest) determined as seeds were used in the rsFC analyses. The seeds were defined as 4mm spheres centered on the peak coordinates of each region from the four proposed emotional regulation networks (ERNs) (Morawetz et al., 2020). The ERN1(A) and ERN2 (B), included 10 and 9 ROIs respectively, are mainly located at fronto-parietal regions and linked to response inhibition, and executive control. While the ERN3 (C) included 8 ROIs are mainly located at subcortical regions and related to memory, emotion perception and generation, the ERN4 (4) included 9 ROIs are mainly related to interoception, body action and awareness.

### Behavior-brain correlation analysis

All subjects in this Neuropsychiatric Phenomics dataset completed measurements of executive function including stop signal task (SST), Balloon analog risk task (BART), and switching task during fMRI scanning, in which fundamental cognitive function comprised of inhibition control and cognitive flexibility were assessed. To explore the correlation between rsFC underlying emotion regulation and cognitive indices, the brain-behavior correlation analyses (Pearson’s correlation) were performed in each group with appropriate multiple comparison correction. In line with previous studies (Lejuez et al., 2002; Verbruggen et al., 2019; Wylie and Allport, 2000), stop-signal reaction time (SSRT), BART risk value and switching costs in reaction time (RT) were calculated respectively as the behavioral indexes.

### Threshold

A cluster-level p < 0.05 FWE (Family Wise Error, FWE; initial thresholding p < 0.001 uncorrected, voxel number≥10) with Bonferroni-correction (p<0.05/36=0.0014) was used for the rest stating functional connectivity analysis. To further disentangle the direction of the group difference and the interaction effect with gender, parameter estimates were extracted and subjected to post-hoc analyses using the significant connectivity areas.

## 3. Results

### rsFC Results

Examining the rsFC underlying emotion regulation between ADHD, BD and HG by means of one-way ANOVAs showed significant FC differences with seeds were mainly located at ERN3 and ERN4. Such as, in the ERN3, the results showed significant group differences for connectivity between left IOG and right lingual/fusiform gyrus (Z = 4.81, p_FWE_<0.001, voxel=165, x/y/z: 18/-74/-8, Fig. 2A). In the ERN4, there was significant group differences between left SPL and Insula/Rolandic operculum connectivity (Z = 4.74, p_FWE_=0.001, voxel=105, x/y/z: -58/-4/12, Fig. 2B) as well as right precuneus and left fusiform connectivity (Z=4.77, p_FWE_=0.001, voxel=105, x/y/z: -30/-82/-16, Fig. 2C). The further post-hoc examination of extracted parameter estimates showed that there was a significantly increased connectivity between left IOG and right lingual/fusiform region in ADHD group compared to both HG and BD group (IOG: HG: p<0.001, BD: p<0.001, Fig. 2A), which indicated an aberrant emotion perception in ADHD than BD and healthy group (Sabatinelli et al., 2011). Similarly, the connectivity between left SPL and insula/Rolandic operculum was significantly increased in ADHD group compared to both HG and BD group (HG: p<0.001, BD: p<0.001, Fig. 2B). In addition, the connectivity between precuneus and fusiform was significantly decreased in both BD and ADHD group compared to HG (BD: p=0.001; ADHD: p<0.001) and there was no significant difference was found between ADHD and BD group (p=0.060; Fig. 2C).

**Fig. 2.**
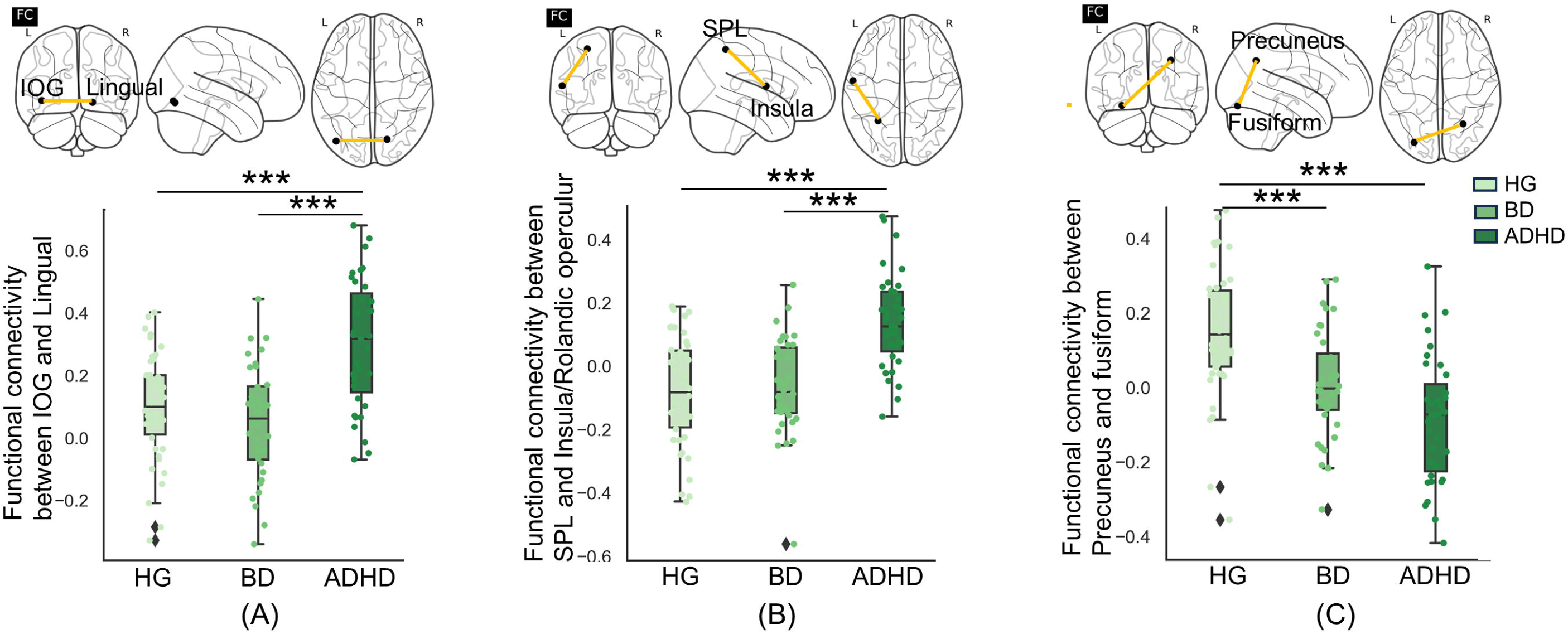
The common and distinct rsFC circuits between ADHD and BD group. The results showed significant group differences for connectivity between (A) left IOG-right lingual/fusiform connectivity and (B) left SPL-Insula/Rolandic operculum connectivity as well as (C) right precuneus and left fusiform connectivity. (A) The further post-hoc examination of extracted parameter estimates showed that there was a significantly increased connectivity between left IOG and right lingual/fusiform region in ADHD group compared to both HG and BD group (IOG: HG: p<0.001, BD: p<0.001). (B) Similarly, the connectivity between left SPL and insula/Rolandic operculum was significantly increased in ADHD group compared to both HG and BD group (HG: p<0.001, BD: p<0.001). (C) In addition, the connectivity between precuneus and fusiform was significantly decreased in both BD and ADHD group compared to HG (BD: p=0.001; ADHD: p<0.001) and there was no significant difference was found between ADHD and BD group (p=0.060).

The exploratory analyses examining the gender effect on the hierarchical functional network between ADHD, BD and HG by means of 2*3 full factor ANOVAS were conducted. The results revealed a significant interaction effect on functional connectivity between IFG/VLPFC (ERN1 ROI) and right supramarginal gyrus (SMG, Z = 3.88, p_FWE_<0.001, voxel=125, x/y/z: 48/-32/34, Fig 3A). In addition, there was a significant interaction effect between left IFG (ERN2 ROI) and ipsilateral DLPFC functional connectivity (Z=4.18, p_FWE_=0.001, voxel=105, x/y/z: -6/32/58, Fig 3B). However, the further post-hoc tests with extracted parameter estimates revealed no significant interaction effect on neither IFG/VLPFC and SMG connectivity (F_(2,107)_=0.139, p =0.871) nor IFG and DLPFC connectivity (F_(2,107)_=1.455, p =0.238).

**Fig. 3.**
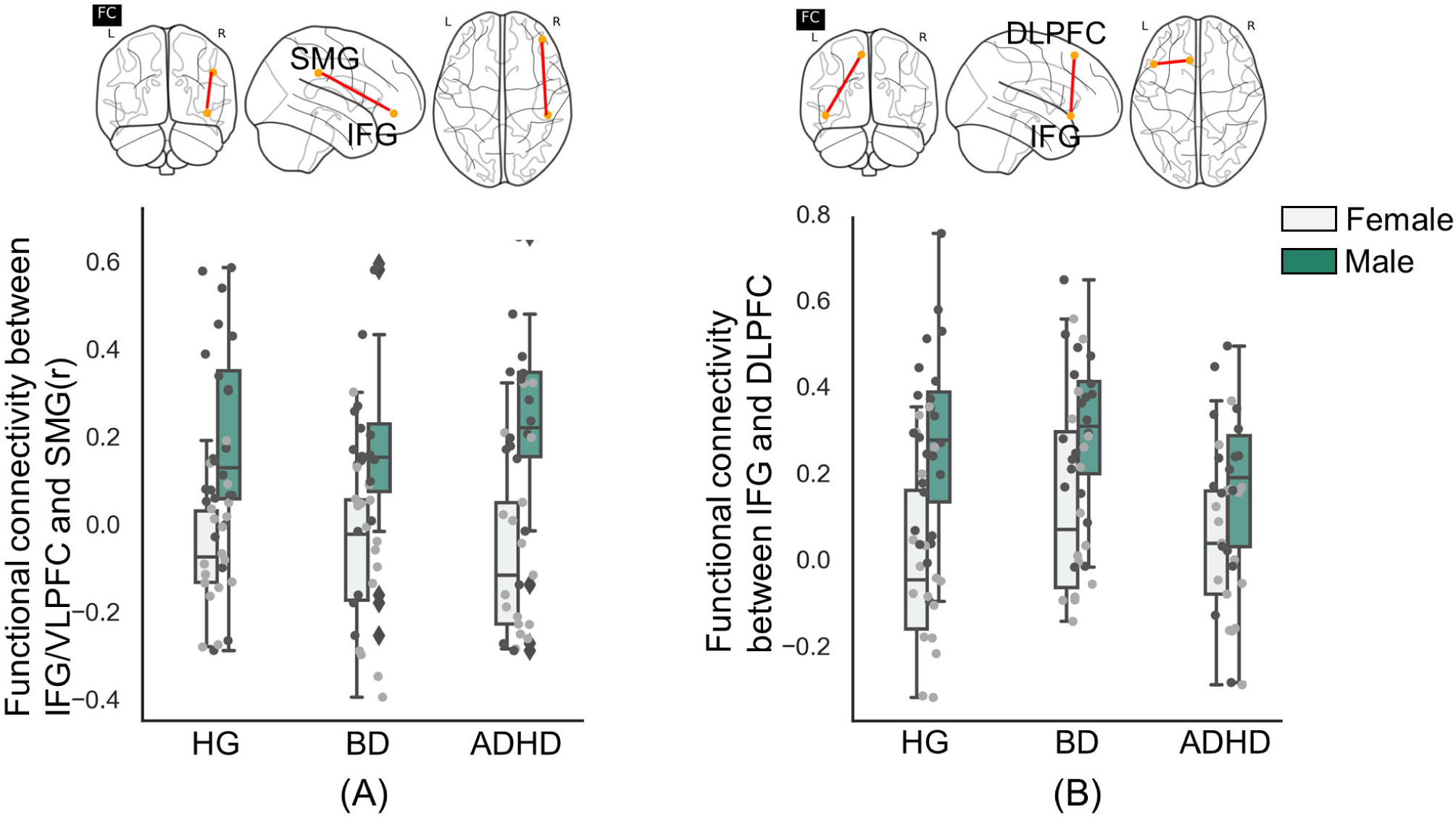
The rsFC results examining the gender and group effect on the hierarchical functional network. The results revealed a significant interaction effect on functional connectivity (A) between IFG/VLPFC and right supramarginal gyrus as well as (B) left IFG and ipsilateral DLPFC. However, the further post-hoc tests with extracted parameter estimates revealed no significant interaction effect on neither IFG/VLPFC and SMG connectivity (F_(2,107)_=0.139, p =0.871) nor IFG and DLPFC connectivity (F_(2,107)_=1.455, p =0.238).

### Association between behavior performance and brain

The association analyses between cognitive indices and functional connectivity revealed no significant correlation between SSRT, BART risk value and switching costs with IOG-lingual, SPL-insula/Rolandic operculum and precuneus-fusiform circuits (SSRT: ps>0.089; BART: ps>0.145; task-switch: ps>0.183).

## 4. Discussion

The current study using the opening dataset sought to explore the common and distinct resting state FC features between ADHD and BD within the framework of hierarchical ER model which included multiple interacting processes of emotion regulation. In generally our findings showed significantly shared and disorder-specific neural circuits implicated in the processes of emotion perception and interception during emotion regulation but not response inhibition or executive control. Such as, the results showed enhanced functional connectivity strength on IOG-lingual/fusiform as well as SPL-insula/Rolandic operculum circuits in ADHD group compared to BD and HG, suggesting impaired emotional facial perception and interoceptive processing in ADHD group. In addition, the connectivity between precuneus and fusiform was significantly decreased in both of ADHD and BD group compared to HG and no significant difference between ADHD and BD group was found. Furthermore, no significant correlation between rsFC strength and cognitive indices in each group were observed. Together these findings can promote the understanding of common and disorder-specific neural mechanism underlying emotion dysregulations and provide some sensitive neural marker to neuroimaging-based clinical evaluation and treatment for ADHD and BD adults.

Our findings focused on the rsFC circuits were in line with previous task-related functional MRI as well as structural MRI studies which showed significant differences between ADHD and BD group on regions or networks related to emotion regulation (Hafeman et al., 2017; Long et al., 2023; Seymour et al., 2015; Seymour et al., 2013). Interestingly, the significant differences between ADHD and BD were only found on neural circuits linking to processes of emotion perception and interception but not response inhibition and executive control. This is in line with previous studies which showing that ADHD and BD group have a comparable response inhibition deficit and emotional impulsivity (Masi et al., 2021; Yep et al., 2018), although no shared neural circuits were found at resting state.

The results reveal an increased functional connectivity between IOG and lingual/fusiform in ADHD group compared to HG and BD group. Given the IOG and lingual/fusiform are key regions engaged into the emotional-related visual information processing, such as early perception of facial features (Furlong et al., 2021; Haxby et al., 2000; Liu et al., 2021), the results suggested impaired emotion perception in ADHD adults group than HG and BD group. This is in line with previous studies which reported that ADHD symptoms are associated with deficits in state and dynamic emotional face recognition (Bozkurt et al., 2024; Uekermann et al., 2010; Zuberer et al., 2022). However, while the recent meta-analysis reported no significant differences on behavioral performance for facial emotion recognition between BD and children and adolescents diagnosed with ADHD (De Prisco et al., 2023), our results revealed enhanced functional connectivity between IOG and lingual/fusiform in ADHD group compared to BD group. This indicates that the differences on the IOG and lingual/fusiform circuits might be a sensitive neural marker to distinguish ADHD and BD group.

We additionally found significantly increased connectivity between SPL and insula/Rolandic operculum in ADHD group than HG and BD group. Based on previous studies showing insula and SPL circuits are mainly associated with interoception and emotional awareness (Jin et al., 2020; Lepping et al., 2015; Simmons et al., 2013), our findings indicated enhanced emotional information processing from internal states in ADHD group compared to HG and BD group. This is consistent with previous study which using resting state data finds that increased clustering and local efficiency of the right insula in the level of functional network was related to the emotion dysregulation in ADHD adults (Viering et al., 2021). Furthermore, we found that the connectivity between precuneus and fusiform was significantly lower in both of ADHD and BD group than HG. Precuneus and fusiform, as the core regions of default mode network (DMN) and visual processing network (VPN), showed altered resting-state functional connectivity as well as decreased grey matter in both ADHD and BD group (Chen et al., 2022; Kucyi et al., 2015; Sutcubasi et al., 2020). More specifically, the intra-network connectivity between DMN and VPN exhibited a strong contribution to emotion classification and sustained emotional experience during naturalistic contexts (Xu et al., 2023). In addition, one previous study has reported that the anti-correlation between precuneus and fusiform were positively related to depression scores (Peng et al., 2015). Thus, the non-significant differences on precuneus and fusiform FC circuits might suggest the shared neural mechanism underlying extensive negative emotion experience between ADHD and BD group.

Our findings showed significant gender and group interaction effect on IFG/VLPFC and right SMG FC circuits as well as left IFG and ipsilateral DLPFC FC circuits which was linked to the processes of response inhibition and executive control, although it failed to be significant in the further post-hoc analysis. Notably, although previous study examining gender effect on subcortical structural morphology between ADHD and BD group has revealed that females in BD group had smaller hippocampal volumes compared to ADHD and HG (Lopez-Larson et al., 2009), studies examining gender differences on rsFC specially associated with emotion regulation remain to be scarce and more future studies could to be conducted.

There are some limitations need to be acknowledged in the current study. Firstly, the participants in the ADHD group included both of predominantly inattentive and hyperactive-impulsive types. Given previous studies have showed that hyperactive-impulsive but not inattentive dimensions are more correlated with emotional dysfunctions in ADHD (Wheeler Maedgen and Carlson, 2000), future studies could be conducted for participants with only hyperactive-impulsive symptoms. Secondly, the brain-cognitive correlation was analyzed with behavioral indexes assessed by SST, BART and switching task which was performed during fMRI scanning. Given the behavioral performance especially the reaction time obtained with fMRI scanning was different from those observed under normal laboratory conditions (Koten et al., 2013), future studies could be conducted to reexamine the association between rsFC and cognitive indexes.

In conclusion, the current study examined the rsFC patterns between ADHD and BD adults within the model of proposed hierarchical ERNs and revealed significantly shared and disorder-specific neural circuits implicated in the processes of emotion perception and interception but not response inhibition or executive control. Such as, we found significantly enhanced functional connectivity strength on IOG and lingual/fusiform as well as SPL and insula circuits in ADHD group compared to HG and BD group. Additionally, the connectivity between precuneus and fusiform was significantly lower in both of ADHD and BD group compared to HG but no significant difference was found between ADHD and BD group. Together these findings can promote the understanding of common and distinguishable neural mechanism linking to emotion dysregulations, which may provide valuable insights into clinical evaluation and treatment.

## CRediT authorship contribution statement

Qian Zhuang: Conceptualization, Formal analysis, Visualization, Writing – original draft; Chuxian Xu: Methodology; Nuo Xu: Formal analysis; Tongtong Wang: Visualization; Matthew Lock: Writing – review; Shuaiyu Chen: Methodology, Supervision

## Declaration of competing interest

The authors declared no conflicts of interest with their research, authorship or the publication of this article.

## Data Availability

The data included in the current study are from online opening dataset(https://www.openfmri.org/dataset/ds000030/)

## Acknowledgments

This work was supported by the National Natural Science Foundation of China (32200904 - Qian Zhuang; 32100888 – Shuaiyu Chen), Zhejiang Natural Science Foundation (LQ21C090007– Shuaiyu Chen) and Medical and Health Technology Project of Zhejiang Provincial Health Commission (ky23023 - Qian Zhuang; 2021ky247– Shuaiyu Chen).

## Notes

### Competing Interest Statement

The authors have declared no competing interest.

### Funding Statement

This work was supported by the National Natural Science Foundation of China (32200904;32100888), Zhejiang Natural Science Foundation (LQ21C090007) and Medical and Health Technology Project of Zhejiang Provincial Health Commission (ky23023; 2021ky247).

### Author Declarations

https://www.openfmri.org/dataset/ds000030/

